# Obesity and longevity: The retrospective longitudinal analysis of international data

**DOI:** 10.1101/2022.06.01.22275879

**Authors:** G. Liapin, V. Vlassov

## Abstract

**Objectives:** Due to the economic growth at the end of the 20th century people in low-income countries live longer, but still spend on food most of their income, and suffer from the low food quality. These countries, as well as affluent countries demonstrate rising prevalence of obesity, which is a risk factor for bad health and increased mortality. The interplay of longevity and obesity at different levels of economic development is not clear.

**Design:** We used World Bank, International Monetary Fund, and Global Burden of Disease project data on population size, longevity, prevalence of obesity, and income as Gross domestic product per capita adjusted to purchasing power in international dollars.

**Results:** After we stratified countries by income, we found that there is a long-term increase in life expectancy with an increase in the prevalence of obesity in countries of all income levels. There is a higher prevalence of obesity in poor countries than in the affluent countries at the equal life expectancy.

**Conclusions:** The economic growth in all countries lead to increase in length of life and is associated with the increase of obesity prevalence. In poor countries the access to food and the food quality is limited as well as customs of consumption are different. It leads to the higher prevalence of overweight and obesity at similar level of length of life. It may be the cause of the slower progress in life expectancy.

## Background

The economic growth at the end of the 20th century made possible deep changes in the consumption of goods and the progress in public health all over the world. People in low-income countries still suffer from hunger, malnutrition and die early, but they live longer life and have more food (1,2). People there fare better, but still spend on food most of their income, and suffer from the low food quality. As a result these countries demonstrates decreasing prevalence of stunting and rising prevalence of obesity (3). In more affluent countries the share of spending on nutrition decreases and shifts towards the consumption of higher quality diverse foods, to savings, travel etc., but the net food consumption is higher (4).

Overweight and obesity appear one of the faces of globalization and urbanization (5,6). Obesity prevalence has doubled since 1980 in more than 70 countries, with a significant growth in other countries (7). Conditions associated with obesity include hypertension, type 2 diabetes, cardiovascular disease, lung disease, and cancer (8,9). It has been shown that obesity has a significant negative impact on life expectancy: severe obesity may be associated with the reduction of life expectancy by 5-20 years (10,11).

The purpose of this study was to analyze the historical connection between the increasing prevalence of obesity and the increase in life expectancy in countries of different income levels.

## Methods

We used data on life expectancy at birth (LEB) and population size from World Bank database (12,13). We used the obesity prevalence data from Global Burden of Disease Project (14) as an age-adjusted prevalence for both sexes, adults (20+ years old). We used the International Monetary Fund Data on GDP purchasing power adjusted in international dollars per capita (GDP PC) (15). All these data were available for 119 countries, listed in the Appendix.

To sort out the effect of the income on the LEB we grouped countries by the GDP PC. We used the average LEB and obesity prevalence in the economically similar groups of countries to smooth the periodic effects of conflicts, crises, and other disasters. We grouped countries by the GDP PC in 1997 into quintiles. Some countries during the study period – 1980-2015 changed their ranking by the GDP PC. To reduce the possible effect of the transition of countries from one quintile of GBP PC to another (e.g. China had moved from lowest quintile to middle of the range), we repeated analysis grouping countries by GDP PC in 1990, 1997, 2005, and 2013. We selected these years to reduce the influence of major world economic fluctuations on the country ranking.

For each group of countries by GDP PC we calculated the weighted average LEB and obesity prevalence for every year. The population size is the weighting factor. Thus, for the group of two countries the weighted average of the prevalence of obesity in the year Y is (pr1Y·pop1Y + pr2Y·pop2Y)/(pop1Y + pop2Y), where pr1 and pr2 are prevalences in these countries, and the pop1 and pop2 are populations in these countries.

For calculations and plotting, we employ Python, libraries pandas and numpy.

This study does not need the approval from ethics review board, because we use publicly available national level data.

## Results

In all five groups by GDP PC there is a strong increase of life expectancy at birth and the prevalence of obesity during period 1980-2015. Fig. 1 presents the results for the case of grouping countries by GDP PC in 1997. When we group countries by GDP PC at the different predetermined time points, we produced similar curves with some irregularities in order of life expectancy curves (see Appendix). Tracks for countries of two upper quintiles of GDP PC are more linear, because the development of member countries was not disturbed by the major natural of social disasters during the study period. Please compare fig. 2, Israel and Argentina as an example.

**Figure 1.**
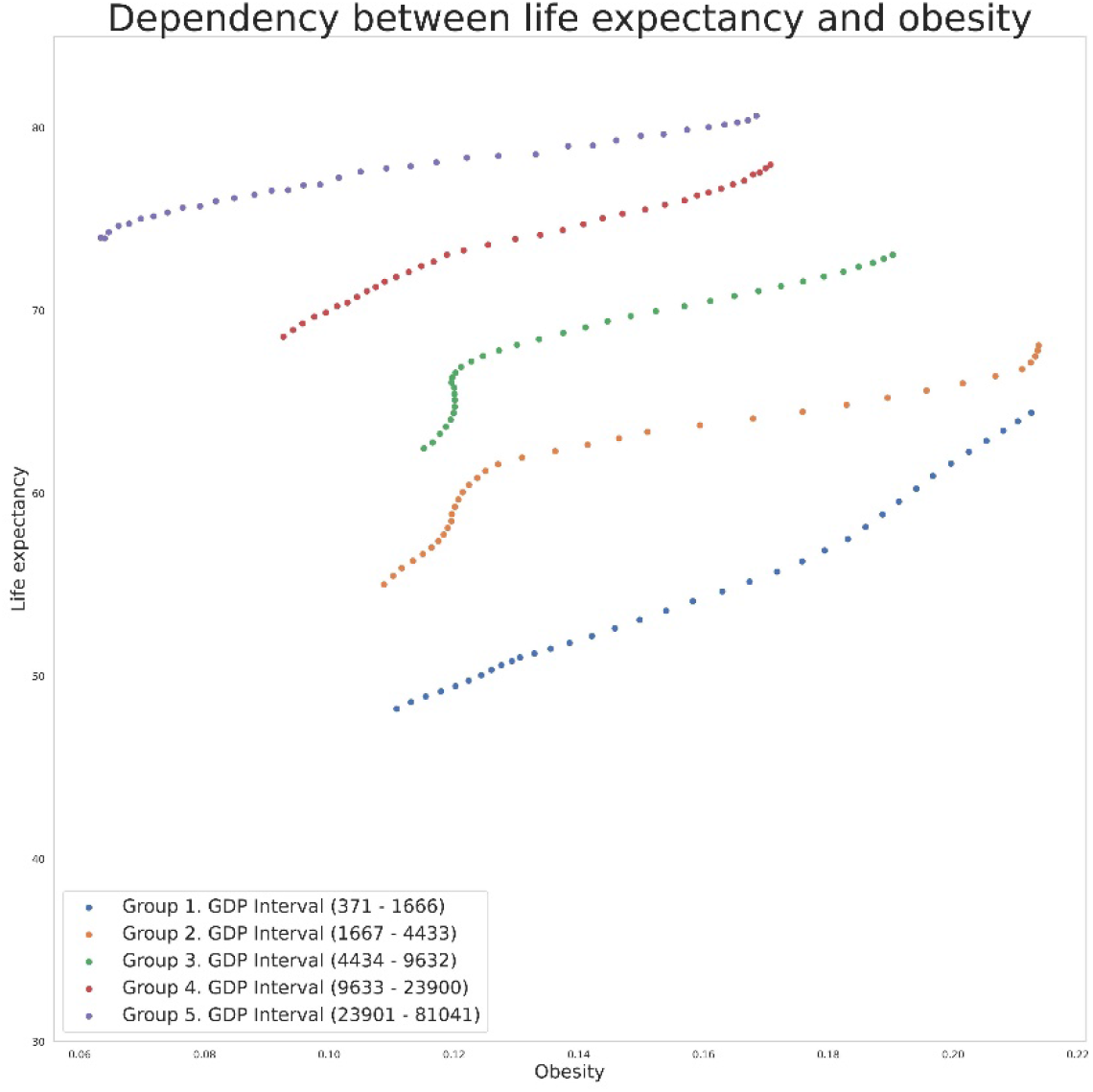
The relationship between LEB and the prevalence of obesity, countries grouped by GDP PC in 1997. Ordinate: LEB, years. Abscissa: Obesity prevalence, %

**Figure 2.**
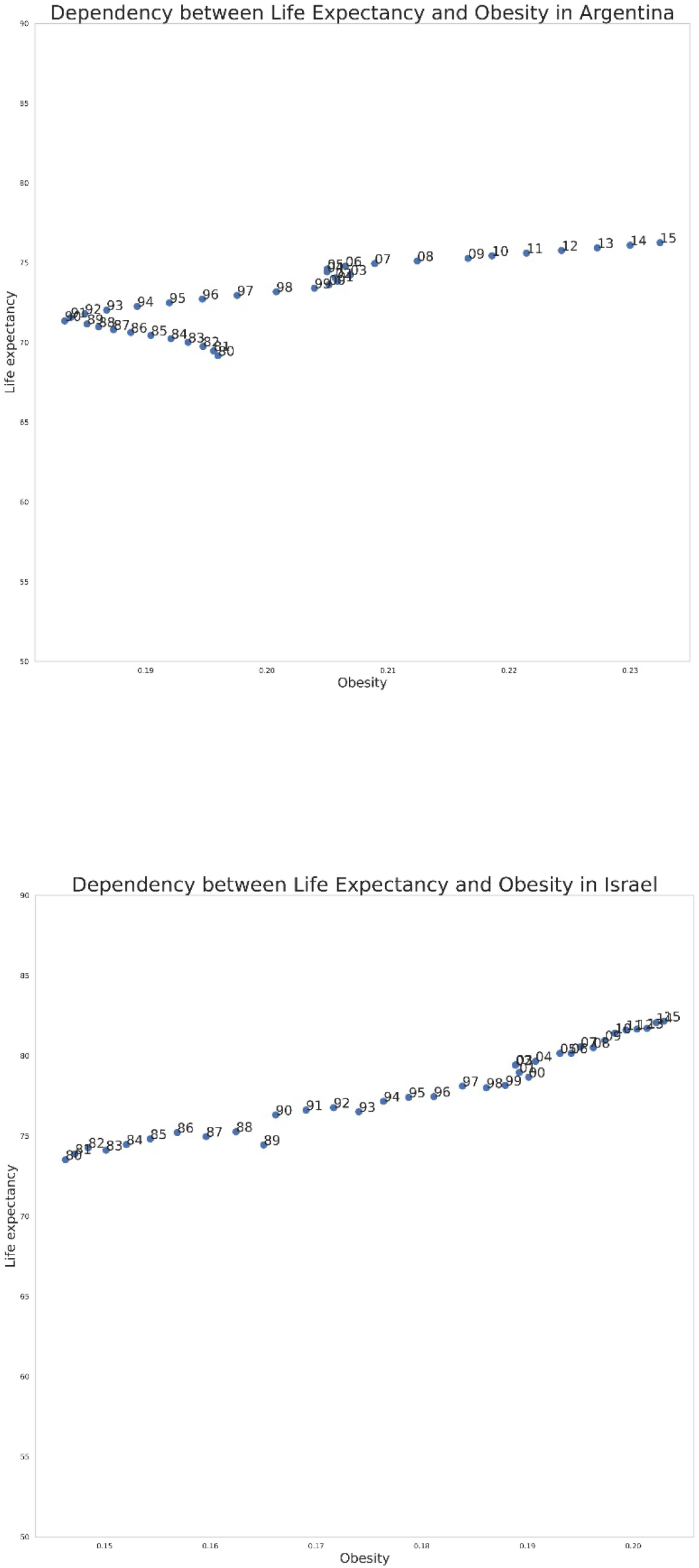
The relationship between the prevalence of obesity (abscissa, %) and LEB in Argentina (a), and Israel (b). Ordinate: LEB, years.

We constructed double charts to examine the additional relations between the three variables (fig. 3). Middle left portion of the chart is similar to the chart at the fig.1. Middle bottom chart is a Preston curve (16). On the logarithmic scale this curve is effectively straightened. Normally the Preston curve is modelled on the surface of scatterplot of the cross-sectional data. Here we see that the average longitudinal data form the same curve.

**Figure 3:**
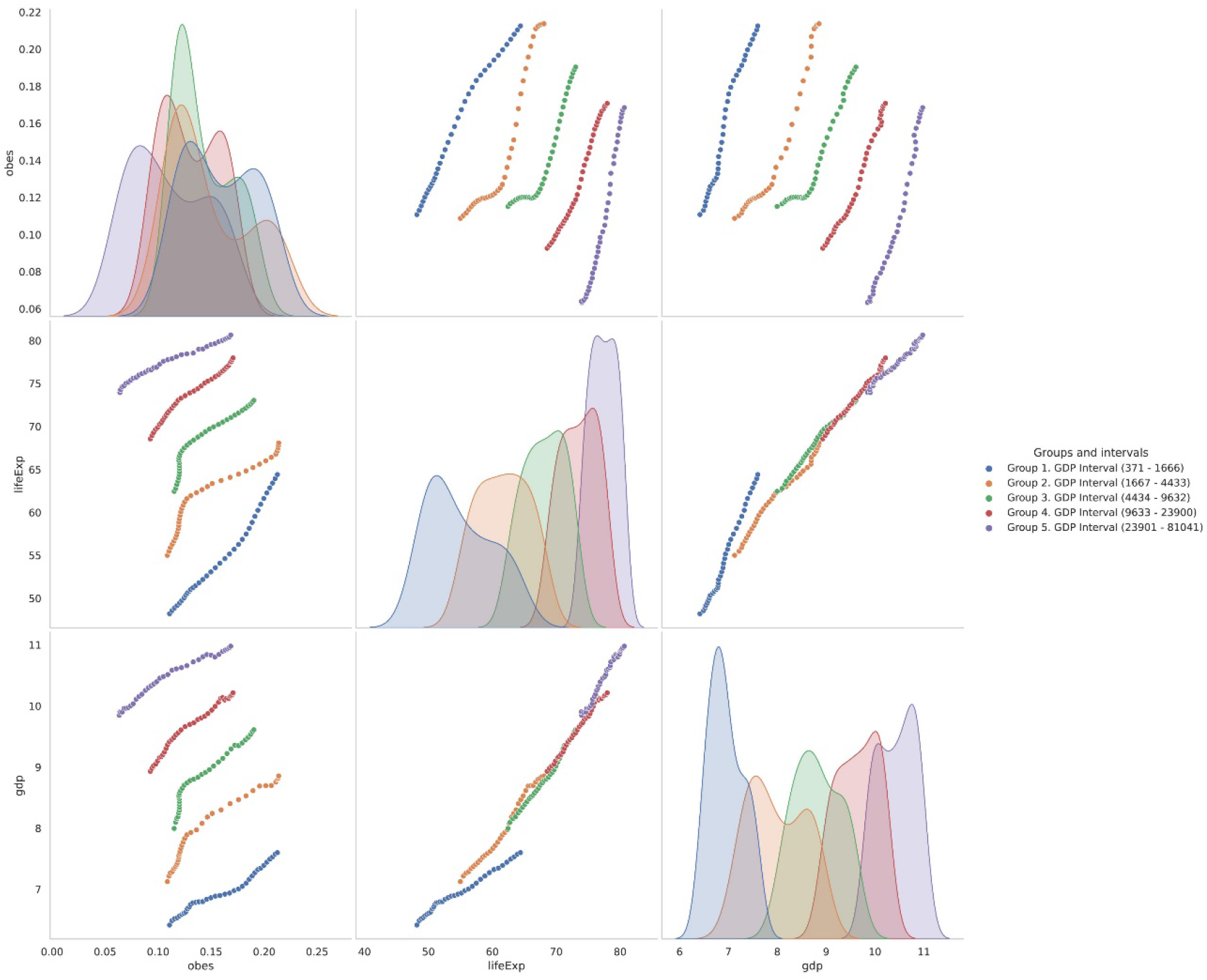
Double chart, grouping of countries by GDP PC 1997. Abscissa: obesity prevalence, %; LEB, years; GDP PC. Ordinate: GDP PC; LEB, years; obesity prevalence (bottom to up). Note log scale for GDP PC

On the fig.2, as well as on the fig. 1, less affluent countries have lower life expectancy and higher prevalence of obesity. The overlap of national data points is significant by the direct comparison, especially of the obesity prevalence data. It is better visible on overlapping histograms of national data points in the five groups. When countries are stratified by GDP PC and pictured longitudinally, the differences are striking.

## Discussion

The main finding of our analysis is that in the all range of countries – from poor to rich – there is a strong historical increase of LEB with growing obesity prevalence. During 35 years of the period of study, the gap between the poor and rich countries had reduced due to faster increase of LEB in poor countries, but the size of the reduction is not appealing. While LEB of poor countries improves, at the similar LEB level the prevalence of obesity is higher in poor countries.

Most research of the influence of overweight and obesity on the longevity are cohort studies on the individual subject level. Our study belongs to the smaller set of research – ecological studies, looking for the differences between populations. Our study compares countries in time. The improvements of the economy and better nutrition necessary should lead to the LEB increase and, in the same time, increase of the obesity prevalence. These very natural parallel changes appear as questioning harms of the obesity. We believe that the explanation is straightforward: socioeconomic development brings so many positive changes to the human life, including better nutrition, housing, education and health care, that it is quite natural that negative impact of the overweight and obesity is negligible comparing to them (17).

Economic growth not just brings the food, but first make accessible the food with cheap calories for the people who never had dreamed about the selection of healthy products. It is the major source of the high and increasing obesity prevalence in developing countries (3,18,19). The low quality of food increasingly available in the poor countries at early stages of the socioeconomic growth may be the explanation of the slow increase of LEB – while the obesity prevalence grows up to 22% (lower curve, fig. 1). Later on, when quality of food and quality of life in general are improving, the growth of longevity became faster. Another force shaping the LEB-obesity curve for the poor countries is the prevalent stunting, leading to the higher body-mass index later in life thus biasing up the estimate of obesity prevalence (20).

In the light of our ecological study it appears, that addressing the high obesity prevalence in the developing countries may be not the primary objective of the public health interventions. The relationships of poor people with the food are very different from the nutritional habits of the people who live in the societies of plenty. Suppression of the excessive calories consumption is rarely possible in poor societies. Only after decades of the good access to the food and appearance of the feeling of safety, the arguments for the healthy food and other possibilities brought by the economic development may be heard and be effective. Imposition of the ideals of the lean body onto the people who in the first family generation have enough food for the day is unreasonable, at least.

Tracks for countries of the two upper quintiles of GDP PC (fig.1) are very similar. The increase of LEB with changes of obesity prevalence is minimal in these groups. It is the probable symptom of the smaller role of nutritional factors in the longevity in the affluent societies. On the other side, in the developed countries the increase of the prevalence of obesity may be a cause of the slow longevity improvement(21,22) and it had been hypothesized that it may be the cause of the future longevity decline (10).

Comparison of the separate countries was not the objective of this study, but examples, offered in fig. 2 are an illustration. The long economic crisis of 1980s in Argentina is reflected in the loop of reduced obesity while slow rise of LEB. The world economic crisis of 1998 led to the depression in Argentina, reflected in the similar, but smaller loop. Another example is Israel, where we can see the short period of reduction of the LEB, which appears the consequence of the economic recession in 1981-1988.

Our study is limited by the nature of data. Many poor countries do not have effective vital registration systems and the estimates of the obesity prevalence are inaccurate or impossible. As a result, the poor countries are underrepresented in our set of countries. In countries included in the study the obesity prevalence data and to a lesser degree longevity data are produced by the relevant organizations using different types of statistical adjustments of field/vital registration data and interpolations. It may lead to the reduction of the random and short-term variations in the individual country data, but may not distort on a large scale the effect we describe.

## Conclusion

During the period 1980-2015 the increase of the longevity at the national level was associated with increase in the prevalence of obesity all around the world. The levels of the past longevity of the affluent countries are attained by low-income countries when they have the higher prevalence of obesity. At the moment the higher longevity is a great achievement, but in future the high prevalence of obesity may be a problem for the developing countries.

## Data Availability

Data used for the analysis are in public domain

## Acknowledgments

We appreciate the support by Semyon Sorokin in calculations and Kristina Rainich for comments on the early draft of the article.

## Web only data

### 1. Countries included in the study

Albania

Algeria

Angola

Antigua and Barbuda

Argentina

Australia

Austria

Bahrain

Bangladesh

Barbados

Belgium

Belize

Benin

Bhutan

Bolivia

Botswana

Brazil

Bulgaria

Burkina Faso

Burundi

Cameroon

Canada

Central African Republic

Chad

Chile

Colombia

Comoros

Costa Rica

Cyprus

Denmark

Dominican Republic

Ecuador

El Salvador

Equatorial Guinea

Ethiopia

Fiji

Finland

France

Gabon

Germany

Ghana

Greece

Grenada

Guatemala

Guinea-Bissau

Guyana

Haiti

Honduras

Hungary

Iceland

India

Indonesia

Ireland

Israel

Italy

Jamaica

Japan

Jordan

Kenya

Kiribati

Kuwait

Lebanon

Lesotho

Libya

Luxembourg

Madagascar

Malawi

Malaysia

Maldives

Mali

Malta

Mauritius

Mexico

Morocco

Mozambique

Nepal

Netherlands

New Zealand

Niger Norway

Oman

Pakistan

Panama

Papua New Guinea

Paraguay

Peru

Philippines

Poland

Portugal

Puerto

Rico Qatar

Romania

Rwanda

Saudi Arabia

Senegal

Seychelles

Sierra Leone

Singapore

Solomon Islands

Spain

Sri Lanka

Sudan

Sweden

Switzerland

Tanzania

Thailand

Togo

Tonga

Trinidad and Tobago

Tunisia

Turkey

Uganda

United Arab Emirates

United Kingdom

United States Uruguay

Vanuatu

Vietnam

Zambia

### 2. Dependence of life expectancy and prevalence of obesity. Countries are grouped by per capita GDP in 1990, 2005, 2013

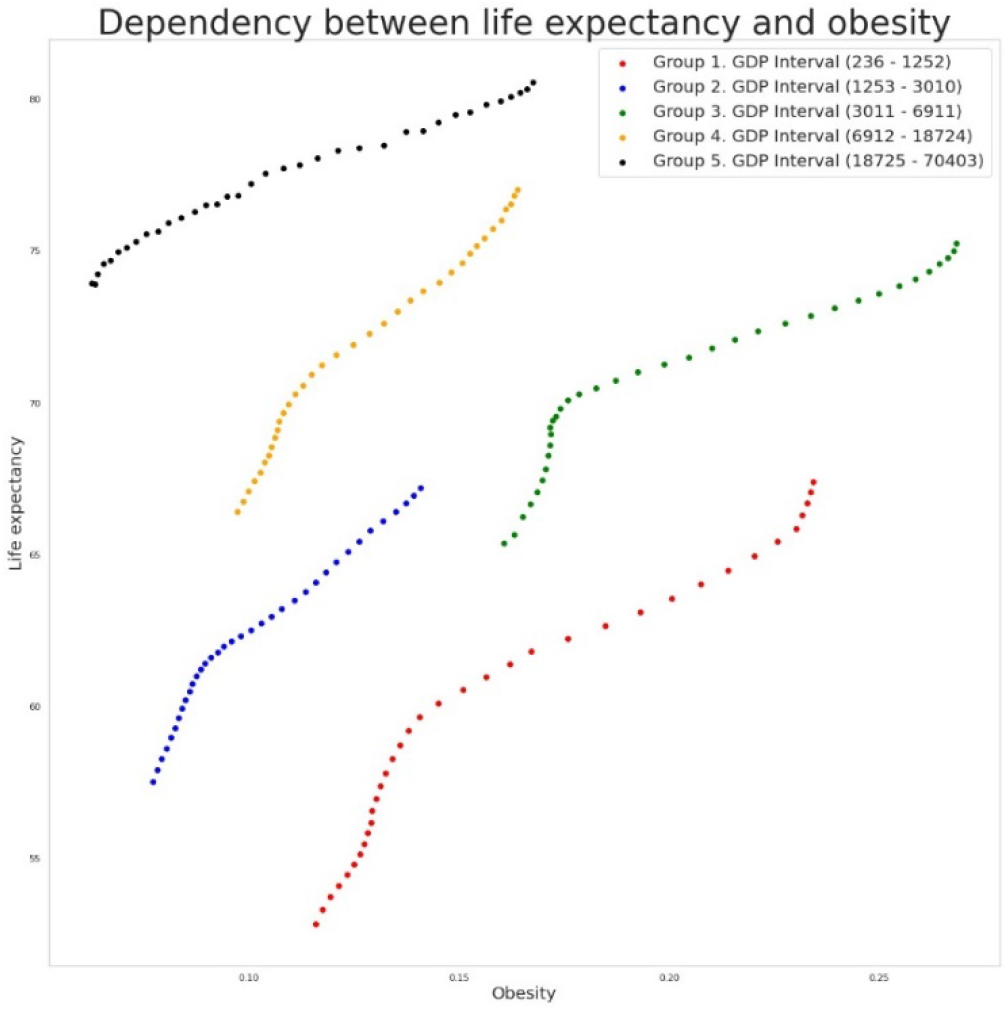

*The relationship between LEB and prevalence of obesity, countries grouped by GDP PC 1990. Ordinate: LEB, years. Abscissa: Obesity prevalence, %*.

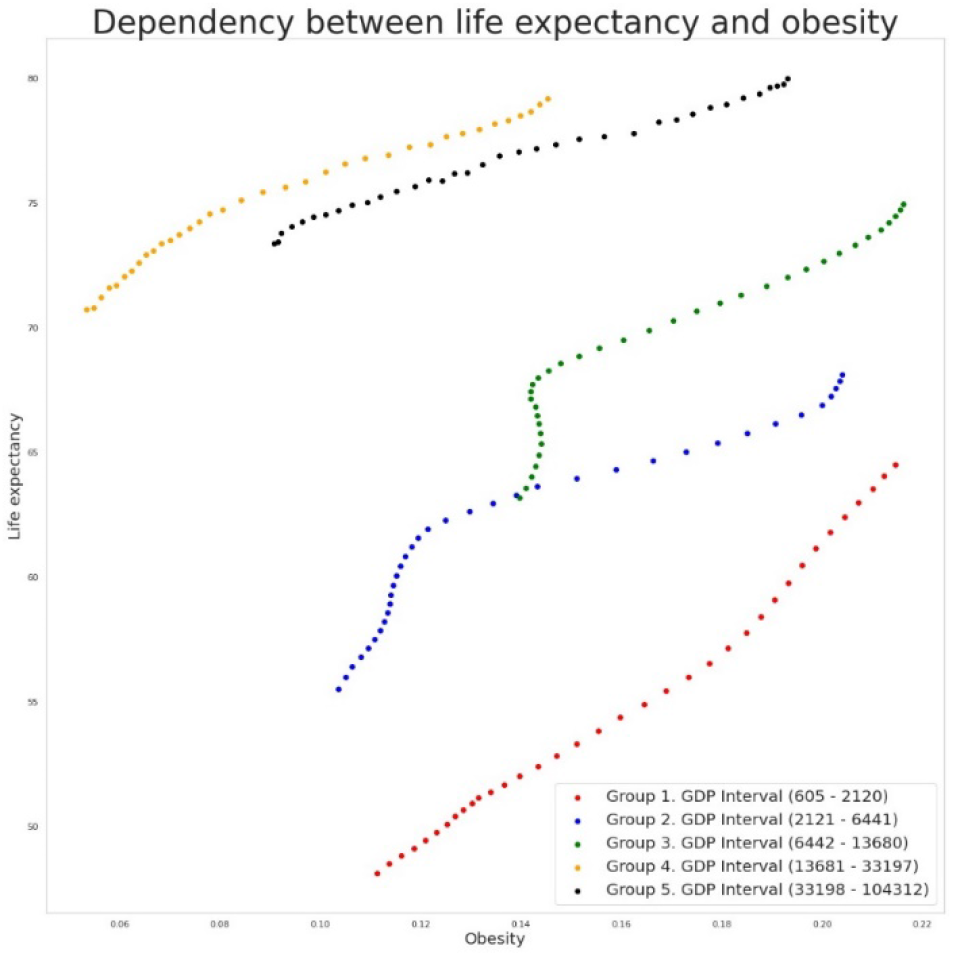

*The relationship between LEB and prevalence of obesity, countries grouped by GDP PC 2005. Ordinate: LEB, years. Abscissa: Obesity prevalence, %*.

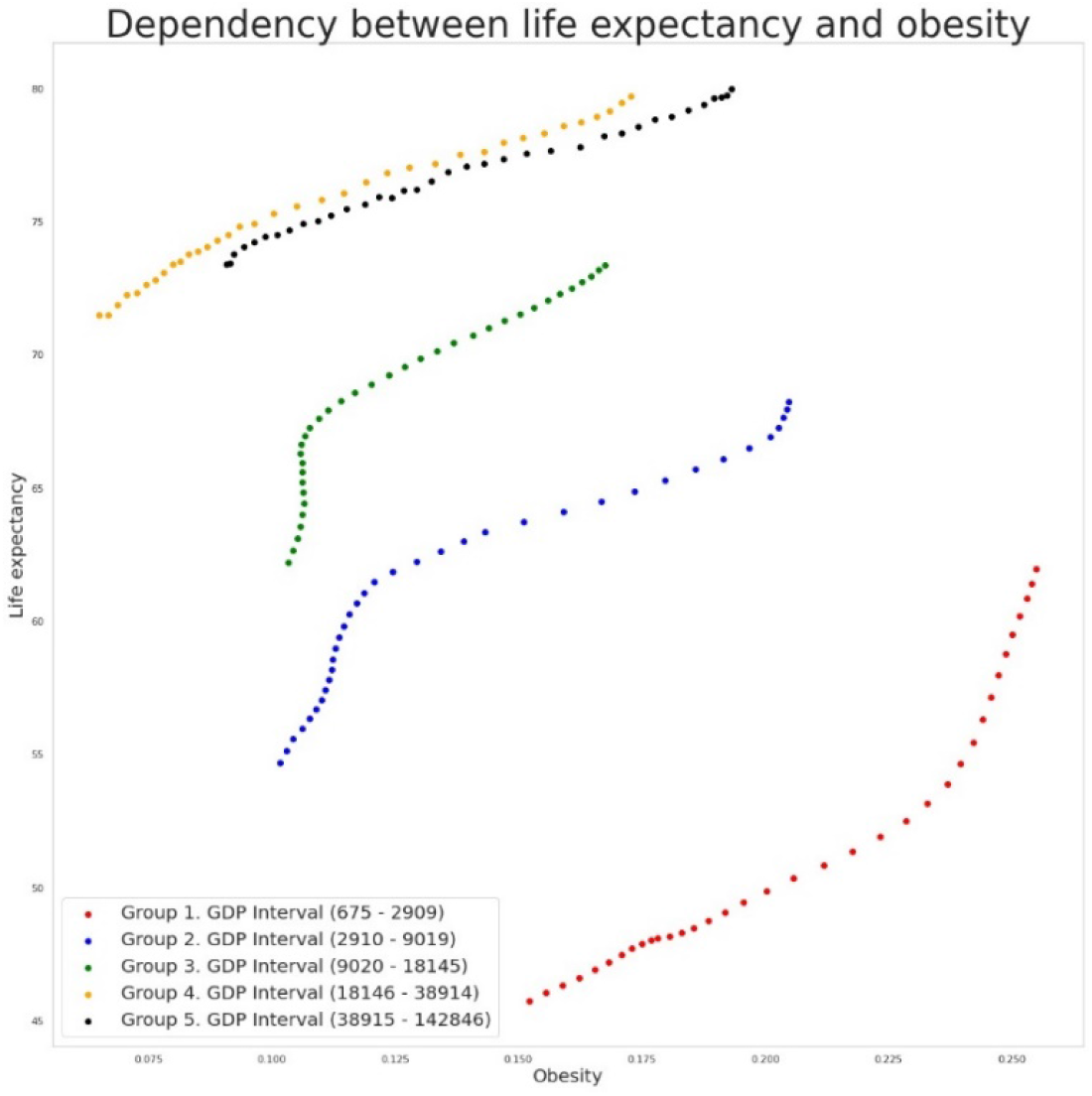

*The relationship between LEB and prevalence of obesity, countries grouped by GDP PC 2013. Ordinate: LEB, years. Abscissa: Obesity prevalence, %*.

